# Ventilation Requirements and Recommendations for Controlling SARS-CoV-2 and Variants Outbreaks in Indoor Gathering Places with Close Contact

**DOI:** 10.1101/2022.06.15.22276447

**Authors:** W.K. Chow

**Affiliations:** Department of Building Environment and Energy Engineering, The Hong Kong Polytechnic University, Hong Kong, China

## Abstract

Unexpected rapid infection involving SARS-CoV-2 variant Omicron known as the fifth wave of outbreak occurred since early January 2022 in Hong Kong. Almost 1.2 million citizens were infected in three months. Ventilation provisions in some gathering places with close contact such as restaurants were found to be lower than requirements, believed to be one of the main causes of transmission in these indoor spaces. At the end of the fifth outbreak in mid-May 2022, group infections were still found in several such gathering places including restaurants and pubs due to inadequate ventilation provisions. There are worries about triggering the sixth wave of outbreak.

Key points related to ventilation requirements in such gathering places are discussed in this paper. Adequate ventilation of 6 air changes per hour minimum must be provided to avoid direct air transmission of virus. Indoor aerodynamics induced by ventilation system must be considered too. However, it is difficult to measure ventilation rate quickly and accurately. A control scheme on virus outbreaks is proposed on installing mechanical ventilation energy use meters and carbon dioxide sensors for checking ventilation provisions adequacy quickly.

## 1. Introduction

Spreading of the novel coronavirus (SARS-CoV-2) and the related disease COVID-19 among people is fast, with over 531 million confirmed cases worldwide already by 9 June 2022 [1]. New SARS-CoV-2 variants such as Omicron are highly infectious. Consequently, people travelling around different places are tightly controlled in some places.

In Hong Kong, those returning from highly infected areas, especially from places with many cases of Omicron or Delta variants, are required to be quarantined for 21 days. This practice has kept zero confirmed infection cases for 7 months up to end 2021. There are some professions exempted from quarantine to keep normal operation of the city. Examples are aircrew and others doing relevant businesses that need frequent travelling. Several such members went out during self-quarantine period and infected others to spread Omicron variant out in a restaurant [2-5]. That is a gathering place with close contact and people taking off their mask. The unexpected fifth wave of infection outbreak was then triggered in January 2022 with up to 1.2 million people infected in 3 months.

After inspecting ventilation provisions and indoor air quality in restaurants with infected cases by government engineers, the overall ventilation rate in some restaurants was below local requirement of 6 air changes per hour (ACH) [2,3,6]. Some areas even had zero air speed and the air filtering system was shut down. This might be the reason why so many customers sitting at different tables were infected. A recent infection case on 22 May 2022 with inadequate ventilation recorded 70 infected cases out of 210 customers in a restaurant [4]. Several other restaurants and pubs are starting to have more customers infected, being a risk of triggering the sixth wave of outbreak. There were also possibilities that some students visiting the identified restaurant and then attended lectures in halls without adequate ventilation provisions. Some students took off the mask while attending lectures as reported in government daily media interview on the virus outbreak.

All these incidents exposed inadequate ventilation provisions to satisfy the local requirement of 6 ACH in those gathering places [2,3,6]. Ventilation rate was marginally under 6 ACH in a restaurant but below 2 ACH in a private party room. Mechanical ventilation system was shut down to get 0 ACH during operation hours in another restaurant during breakfast time. This raised the issue of controlling and inspecting ventilation provisions quickly at gathering places including catering areas, lecture halls of education institutes and public transport vehicles. Adequate ventilation must be provided in restaurants because people take off their masks while eating and drinking. It is understood that electricity bill for running ventilation system is very high. This part will be discussed later with recommendations made.

## 2. Simple Modeling

Most of the current ventilation design guides follow the theory based on a well-mixed model [7]. In a room with space volume V, the fresh air intake rate and the exhaust rate are taken to be the same, at a value of N_A_ (expressed in ACH). Sources of pollutant are taken to be of emitting rate *α* (expressed in normalized source strength), giving an instantaneous pollutant or dust concentration of C (expressed in kg per kg of air) at time t. The mass balance equation for thorough mixing of pollutant and air in a well-mixed model [7] gives:

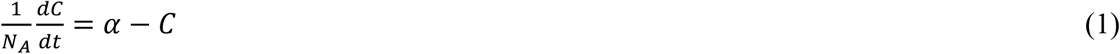

Different values of α would give different mass balance equations [7] for solving the pollutant concentration. If the value of α is a constant, direct integration of equation (1) gives transient pollutant concentration. This equation is used to study pollutant or dust concenteation. The ventilation requirement expressed in terms of *N*_*A*_ was worked out based on acceptable carbon dioxide level for general indoor ventilation and carbon monoxide concentration for car parks or vehicular tunnels.

Ventilation provided for COVID-19 virus control was studied in a slighty different way. A large amount of viruses is unlikely to survive in air without substrate and aerosol. Air transmission of COVID-19 is generally believed to be caused by virus on droplets from coughing and other expiratory activities [8-10]. Indoor virus transmission through air can be controlled by providing adequate ventilation. Increasing *N*_*A*_ would improve the indoor air quality.

Effect of *N*_*A*_ [11-14] on virus transmission rate of rooms was investigated. The Wells-Riley (WR) infection model [15,16] with the probability *P* of infection risk for a susceptible can be simplified under appropriate conditions in terms of *N*_A_ through a parameter *A* with limited experimental data available as:

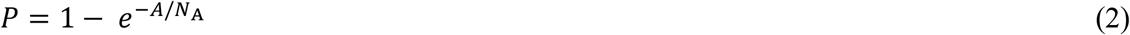

There are very few updated data for the novel coronavirus SARS-CoV-2, especially for the more infectious mutant coronavirus strains. Based on available literature experimental results, *P* was fitted [16] against *N*_A_ with Eq. (2) to yield *A* = 0.8054 hr^-1^, with a low correlation coefficient of 0.4621.

Values of *P* varied largely for *N*_A_ less than around 6 ACH [16]. This suggested that *N*_A_ should be over 6 ACH to give a low enough *P*.

## 3. Ventilation Requirements

A 6 ACH is required for COVID-19 spread control in gathering places including restaurants [2,3]. However, small VIP rooms of restaurants had inadequate ventilation provision of 2 ACH instead of 6 ACH as observed in the fifth wave of outbreak, some even had 0 ACH in that eatery for breakfast! Low ventilation rate will have the risk of transmitting virus within a short time.

Direct air transmission was suspected [17] to be possible in a hotel room first. In the outbreak of virus variant in the restaurant concerned, the ventilation rate was less than 6 ACH, only 2 ACH in an isolated room [4,5,18-23]. However, transmission of COVID-19 directly through air had not yet been confirmed. If virus can transmit through air directly, not just several customers should have been infected. As reported in the above literature [8-10], the size of virus is small and has to deposit on fine dust or very fine droplets (to give aerosols), then transmission by air through mixing. Such infected fine dust are not distributed uniformly in the restaurant.

Safety culture [24-26] is important to ensure that safety management works accordingly. There are always conflicts between safety provisions and environmental protection. Safety, thermal comfort and building energy use have to be compromised accordingly. It is understood that decision is difficult to make, always having challenges on handling management issues.

Ventilation provisions to some buildings with central air-conditioning systems were turned off outside office hours for energy conservation. This would expose staff working early or late at night to high risk. Ventilation operation schedule in workspace must match with opening time, though building energy use is another concern.

There are even doubts on some university lecture rooms having inadequate ventilation. University management must ensure that ventilation provisions comply at least with the required specifications of 6 ACH. They must watch carefully because they are educating young citizens in shouldering responsibility.

Inadequate ventilation provision in public transport such as light buses run by small and medium enterprises (SME) and even buses were discussed [27]. Virus can be transmitted easily among passengers in public light bus for 21 people. The ventilation system might be occasionally shut off to save some operation cost. It is very difficult to inspect whether ventilation provision is 6 ACH at the moment. Mandatory installation of an energy use meter and a carbon dioxide meter might be required to monitor key parameters. Transient carbon dioxide concentrations [28,29] measured can then be used to deduce ventilation rate from equation (1) derived from the well-mixed model in a later section.

## 4. Indoor Aerodynamics

As reported on studying building ventilation requirements in 2000 [16], indoor air flow pattern should be studied carefully for developing codes related to ventilation requirements for the catering services. Air flow pattern is important in the mixing of clean and polluted air. Local air speeds and turbulence at occupied zones are the key factors determining indoor environmental quality [30-44]. However, putting partitions between tables might block air flow and promote direct air transmission [32] in those stagnant positions. Increase in turbulence intensity will give better mixing of air with contaminants. Computational Fluid Dynamics (CFD) is now commonly used to study the air flow and turbulent mixing [33,41]. CFD is relatively cheaper than other methods in physical experimental study [32].

In areas with high humidity, both the number and size of water droplets increase [12-15]. There is more water surface for more virus to stay. Infection transmission rate should be higher. But under dry environment, droplets should be evaporated faster. Very fine droplets can even be evaporated completely. If the environment is very clean with low concentration of solid particulates, small SARS-CoV-2 virus of 50 nm to 200 nm has not so much substrate surface area to deposit when the liquid droplets completely evaporate. The disease transmission rate should be reduced as observed in an aircraft cabin with dry and clean environment with low particles concentration [14].

## 5. Control Scheme

Inadequate ventilation provisions (below agreed values such as 6 ACH) in gathering places such as catering areas, university lecture halls and public transport vehicles would have higher virus transmission rate. It is very difficult to measure the ventilation provisions in a big space quickly with acceptable accuracy.

Earlier observations on ventilation inadequacy in a train car with passengers trapped inside [28,29] suggested that carbon dioxide concentration can give an estimation on ventilation rate based on the well-mixed model equation (1). Carbon dioxide concentration will be very high for inadequate ventilation rate. Based on this observation, the following are suggested along two lines:

- Measuring transient concentration of carbon dioxide at appropriate positions determined by indoor aerodynamics pattern can estimate the instantaneous ventilation rate based on the well-mixed model in a suitable region [43]. These points can be determined by field measurement or CFD.
- Installing an energy use meter [29,40] to the mechanical ventilation system will also give supporting data for checking whether the system is operating according to the ventilation provision strategy.

This control strategy should be considered by putting in the facility management office on disaster management, similar to ensure fire safety management as pointed out to be under facility management [45]. Energy use impact for ventilation in controlling virus transmission through air should be handled properly with further studies [46].

## 6. Conclusions

Hong Kong kept the number of local cases at zero for 7 months till end 2021 in blocking outsiders around. However, the fifth wave of outbreak led to 1.2 million citizens infected in three months. Waiving quarantine in designated monitored places becomes a loophole in controlling infection from outside high-risk areas with huge number of cases of infection. Some members did not follow agreed relaxed quarantine by home isolation and joined lunch gathering at restaurant with mask taken off while dining, visited shopping malls and took public transportation during holidays. Even those customers sitting at the restaurant entrance were affected. Consequently, their lunch partners were infected in those spaces without adequate ventilation. Authorities then have tightened regulations over some professionals with more relaxed quarantine period.

A value of 6 ACH is required [2,3] specifically for COVID-19 spread control in many building uses including catering services, university lecture rooms and public transport vehicles including public light buses run by SME. This adopted value appears to be effective but has led to many challenges. Operating cost for mechanical ventilation system is very high. That might be the reason why some places have 0 ACH. Flow characteristics in mechanical air handling system take a longer time to measure. It is not easy to inspect quickly whether the ventilation rate is 6 ACH. It is even more difficult to check whether the ventilation system is occasionally shut off to save some fuel cost. Virus can then be transmitted easily among passengers.

Key safety management schemes for controlling virus outbreak in those indoor gathering places with close contact are:

- Ventilation provisions to those gathering places including catering services, university lecture halls and public transport must follow the regulations of keeping a minimum of 6 ACH.
- Whether direct air transmission of the new virus variants is possible should be further explored.
- Reducing operation cost by lowering electricity price, and waiving tax for fuel in public transport system.
- It is very difficult to inspect whether ventilation provision is 6 ACH quickly and accurately. Mandatory installation of an energy use meter and carbon dioxide sensors at appropriate positions might be required for quick inspection of the ventilation rate.

## Supporting information

ICMJE DISCLOSURE FORM, and will be used for the link to the file on the preprint site.

## Data Availability

All data produced in the present work are contained in the manuscript

